# Medication, Vaccine, and Folic Acid Use Among Pregnant Women in Belgium: Insights from the BELpREG Cohort

**DOI:** 10.64898/2026.02.04.26345552

**Authors:** Laure Sillis, Sien Lenie, Emily Jacobs, Karel Allegaert, Annick Bogaerts, Maarten De Vos, Titia Hompes, Anne Smits, Kristel Van Calsteren, Jan Y Verbakel, Veerle Foulon, Michael Ceulemans

## Abstract

**Background:** Safety data for most medications in pregnancy remain limited, yet pharmacological treatment is often necessary. Evidence on real-world medication use in pregnancy including over-the-counter products and folic acid is scarce, especially in Belgium.

**Methods:** We conducted a drug utilisation study using self-reported data from BELpREG, a prospective, web-based pregnancy registry established in November 2022. Pregnant individuals aged ≥18 years receiving healthcare in Belgium can enrol voluntarily at any stage in pregnancy and complete online questionnaires at enrolment and every four weeks until delivery. All participants with follow-up beyond the first trimester were included, and trimester-specific cohorts were constructed based on completion of questionnaires after each trimester. Data were extracted in July 2025.

**Results:** This study included 2,096 participants, of whom 1,767 were followed through trimester 2 and 1,136 through trimester 3. Median gestational age at enrolment was 16 weeks. Prevalence estimates of medication use were 80.2% in the six months before conception, 85.8% in trimester 1, 92.0% in trimester 2, and 94.9% in trimester 3. The most common classes were analgesics, vaccines, antihistamines, antianemic preparations, and drugs for acid-related disorders. Paracetamol was most frequently used (35.4% in trimester 1), typically short term (median 3 days), followed by doxylamine-pyridoxine (26.7% in trimester 1). Folic acid supplementation was nearly universal, though only 59.9% met national guideline-concordant criteria. Maternal vaccine uptake was substantial but incomplete, with 67.2% receiving pertussis, 41.5% influenza, and 21.5% COVID-19 vaccination. Exposure to potentially inappropriate or teratogenic medications was rare.

**Conclusions:** Medication use during pregnancy in Belgium was nearly universal, with high use of paracetamol and doxylamine-pyridoxine. Folic acid and vaccine uptake were substantial, but often not guideline-concordant.

**Key Points:**

- Medication use during pregnancy in Belgium was nearly universal, with over 85% of participants reporting use in the first trimester and 95% in the third.
- Paracetamol (35% in the first trimester) and doxylamine–pyridoxine (27% in the first trimester) were the most frequently used medications.
- Folic acid use was widespread, yet only about 60% of participants followed national timing and duration recommendations.
- Maternal vaccine uptake was substantial, particularly for pertussis (67%), though not universal despite guideline recommendations
- BELpREG’s self-reported data capture both prescription and over-the-counter medications, offering a complete picture of real-world use during pregnancy.

**Plain Language Summary:** This study looked at how often and when people in Belgium use medications, folic acid supplements, and vaccines during pregnancy. Using data from the BELpREG pregnancy registry, more than 2,000 pregnant participants completed online questionnaires about their health and medication use throughout pregnancy (every four weeks). Almost everyone reported taking at least one medication: 86% during the first trimester and 95% during the third. The most common medicines were paracetamol and doxylamine–pyridoxine. Nearly all participants used folic acid, but only about 60% followed national recommendations for starting timely before pregnancy and continuing through the first trimester. Many received recommended vaccines during pregnancy: about 67% for pertussis, 42% for influenza, and 22% for COVID-19; but uptake was still incomplete. Exposure to potentially inappropriate or teratogenic medications was rare. Because BELpREG collects self-reported data, including both prescribed and over-the-counter products, it provides a comprehensive picture of real-world medication use in pregnant people. Further, these findings help identify gaps between guideline recommendations and actual practice.

**Social Media Quote:** Medication use in pregnancy is nearly universal in Belgium. Paracetamol and doxylamine-pyridoxine top the list. Folic acid and vaccine uptake are high but often not guideline-concordant. BELpREG data reveal unique self-reported real-world patterns. #BELpREG

## 1. Introduction

Not all medications are safe to use during pregnancy. Some prenatal exposures increase the risk of adverse outcomes such as congenital anomalies, spontaneous abortion, preterm birth, and impaired child neurodevelopment. Yet, safety data for most medications remain limited [1, 2], largely due to the exclusion of pregnant women from clinical trials [3, 4]. At the same time, pharmacological treatment for chronic or pregnancy-related conditions is often indispensable, making medication use during pregnancy common and clinically warranted.

Internationally, more than 80% of pregnant women in high-income countries report medication use, though prevalence varies by population and data source [5-8]. Frequently reported classes include analgesics, anti-infectives, antacids, antiemetics, and antihistamines, with substantial inter-regional variability [6]. Several studies also document the use of potentially harmful or teratogenic agents [5, 9-11], underscoring the need for enhanced monitoring and counselling.

In Belgium, epidemiological data on actual medication use during pregnancy are scarce. A 2019 cross-sectional study at a tertiary hospital showed frequent use of medication and supplements, but covered only a seven day recall window, thus missing exposures across gestation [12]. Nationwide dispensing data from the BeMeP database (2010– 2016) revealed population-level trends [13] but excluded over-the-counter (OTC) products and lacked precise gestational timing, preventing accurate attribution of exposure to pregnancy. These limitations underscore the need for complementary, prospective data reflecting real-world, self-reported medication use in pregnancy.

To fill this data gap, the BELpREG pregnancy registry was launched in Belgium in November 2022 (www.belpreg.be/en) [14, 15]. BELpREG is an ongoing, prospective, web-based registry that was primarily established to evaluate medication and health products’ safety during pregnancy. To achieve this, BELpREG collects detailed longitudinal, self-reported information on all medication exposures, alongside vaccines, supplements, and other health products. Pregnant individuals aged ≥18 years receiving healthcare in Belgium can voluntarily enrol at any stage of pregnancy. After electronic consent, participants complete an enrolment questionnaire and follow-ups every four weeks until delivery, with postpartum questionnaires extending to four years of infant’s age. Recruitment is carried out through multiple channels, including healthcare providers and social media.

## 2. Objectives

The four-weekly questionnaire design of BELpREG enables detailed, time-specific assessment of medication use, capturing both prescription and OTC products. This study examined the prevalence of medication use during pregnancy in Belgium across all therapeutic classes and by timing of exposure, while also documenting folic acid intake and vaccination patterns. Together, these findings provide a comprehensive picture of real-world medication use in pregnancy, highlighting commonly used substances with limited safety evidence, and identifying non-optimal use patterns that can guide public health policy and clinical counselling.

## 3. Methods

This observational drug utilization study was conducted using data from the BELpREG pregnancy registry, a prospective, web-based registry designed to collect longitudinal, self-reported information on medication, vaccine, and folic acid use during and around pregnancy in Belgium [14, 15]. Ethical approval was granted by the Ethics Committee Research UZ/KU Leuven (S66464), and all participants provided electronic informed consent.

### Study population

The study population comprised all BELpREG participants who completed the enrolment questionnaire, up until the questionnaire assessing medication use in the six months preceding conception, and who remained in follow-up in BELpREG beyond the end of the first trimester of pregnancy. For trimester-specific analyses, participants were assigned to nested cohorts if they had completed the medication questionnaire after the end of the relevant trimester. Thus, a participant could contribute to multiple trimester cohorts. Pregnancies were included regardless of outcome status at the time of data extraction (July 2025).

### Data collection

Medication, vaccine, folic acid, and other health products were reported via structured online questionnaires at enrolment, every four weeks during pregnancy, and once in the early postpartum period (within four weeks after delivery). Enrolment was possible at any stage of pregnancy, with the enrolment questionnaire covering exposures since conception and preconception use. Each follow-up questionnaire assessed exposures since the previous one; missed questionnaires triggered a reminder after 14 days and did not prevent subsequent follow-ups. REDCap software was used for data collection. Survey instruments were linked to two national resources: the official Belgian medication database (Source Authentique des Médicaments, SAM) and the Medipim database, which provides images of product packaging. Participants selected products from structured drop-down menus; images were displayed to enhance accurate reporting. A detailed overview of the medication, vaccine, and folic acid use-related questions in BELpREG is provided in the Supplementary Table 1.

**Table 1.** Self-reported characteristics of BELpREG participants in the nested pregnancy trimester cohorts.

|  | Follow-up until the end of trimester 1 (N=2096) |  | Follow-up until the end of trimester 2 (N=1767) |  | Follow-up until the end of trimester 3 (N=1136) |  |
| --- | --- | --- | --- | --- | --- | --- |
|  | n | % | n | % | n | % |
| Language of completing the questionnaires |  |  |  |  |  |  |
| Dutch | 1914 | 91.3 | 1613 | 91.0 | 1040 | 91.5 |
| French | 159 | 7.6 | 134 | 7.6 | 83 | 7.3 |
| English | 23 | 1.1 | 20 | 1.1 | 13 | 1.1 |
| Maternal age at LMP |  |  |  |  |  |  |
| <25 years | 41 | 2.0 | 30 | 1.7 | 20 | 1.8 |
| 18-29 years | 648 | 30.9 | 554 | 31.4 | 353 | 31.1 |
| 30-34 years | 1078 | 51.4 | 911 | 51.6 | 598 | 52.6 |
| 35-39 years | 287 | 13.7 | 237 | 13.4 | 142 | 12.5 |
| >40 years | 37 | 1.8 | 32 | 1.8 | 21 | 1.8 |
| Missing | 5 | 0.2 | 3 | 0.2 | 2 | 0.2 |
| Maternal ethnical background |  |  |  |  |  |  |
| Caucasian | 2062 | 98.4 | 1743 | 98.6 | 1122 | 98.8 |
| Non-Caucasian | 34 | 1.6 | 24 | 1.4 | 14 | 1.2 |
| Civil status |  |  |  |  |  |  |
| Partner | 2045 | 97.6 | 1724 | 97.6 | 1113 | 98.0 |
| No partner | 51 | 2.4 | 43 | 2.4 | 23 | 2.0 |
| Maternal education |  |  |  |  |  |  |
| Low/ medium | 278 | 13.3 | 223 | 12.6 | 133 | 11.7 |
| High<sup>1</sup> | 1818 | 86.7 | 1544 | 87.4 | 1003 | 88.3 |
| Maternal employment status in the past year |  |  |  |  |  |  |
| Employed | 2039 | 97.3 | 1719 | 97.3 | 1107 | 97.4 |
| Not employed | 57 | 2.7 | 48 | 2.7 | 29 | 2.6 |
| Annual household gross income |  |  |  |  |  |  |
| <45,000 euros | 248 | 11.8 | 205 | 11.6 | 128 | 11.3 |
| 45,000 – 65,000 euros | 477 | 22.8 | 412 | 23.3 | 262 | 23.1 |
| >65,000 euros | 856 | 40.8 | 717 | 40.6 | 475 | 41.8 |
| I don't know/ I'd rather not tell | 515 | 24.6 | 433 | 24.5 | 271 | 23.9 |
| Gravidity |  |  |  |  |  |  |
| Primigravida | 962 | 45.9 | 823 | 46.6 | 541 | 47.6 |
| Multigravida | 1134 | 54.1 | 944 | 53.4 | 595 | 52.4 |
| Method of conception |  |  |  |  |  |  |
| Spontaneous | 1724 | 82.3 | 1464 | 82.9 | 943 | 83.0 |
| ART | 372 | 17.7 | 303 | 17.1 | 193 | 17.0 |
| Multiple gestation<sup>2</sup> |  |  |  |  |  |  |
| Singleton pregnancy | 1853 | 88.4 | 1586 | 89.8 | 1036 | 91.2 |
| Multiple pregnancy | 21 | 1.0 | 17 | 1.0 | 4 | 0.4 |
| I don't know yet | 222 | 10.6 | 164 | 9.3 | 96 | 8.5 |
| Planned pregnancy<sup>3</sup> |  |  |  |  |  |  |
| Planned | 1924 | 91.8 | 1629 | 92.2 | 1053 | 92.7 |
| Unplanned | 172 | 8.2 | 138 | 7.8 | 83 | 7.3 |
| Any chronic condition prior to pregnancy<sup>4</sup> | 841 | 40.1 | 707 | 40.0 | 448 | 39.4 |
| Allergic & Hypersensitivity Conditions | 222 | 10.6 | 195 | 11.0 | 125 | 11.0 |
| Immune-mediated Inflammatory diseases | 184 | 8.8 | 150 | 8.5 | 94 | 8.3 |
| Thyroid Disorders | 130 | 6.2 | 113 | 6.4 | 76 | 6.7 |
| Gastrointestinal Diseases | 120 | 5.7 | 101 | 5.7 | 65 | 5.7 |
| Asthma | 92 | 4.4 | 82 | 4.6 | 55 | 4.8 |
| Diabetes & Insulin Resistance | 40 | 1.9 | 38 | 2.2 | 16 | 1.4 |
| Depression and Depressive Disorders | 26 | 1.2 | 24 | 1.4 | 13 | 1.1 |
| Other Mental Health Disorders<sup>5</sup> | 60 | 2.9 | 45 | 2.6 | 33 | 2.9 |

All results are shown as absolute numbers (n) and percentages (%); Missing data, if applicable, are indicated for each variable in the table; ART: assisted reproductive technology; <sup>1</sup>High education is defined as any formal degree obtained in higher education or university; <sup>2</sup>Asked at enrolment; <sup>3</sup>A planned pregnancy is self-defined as a yes-no question at enrolment; <sup>4</sup>Chronic conditions that were present (and known) prior to the onset of the current pregnancy were self-reported by BELpREG participants during the enrolment questionnaire; <sup>5</sup>Including addiction, agoraphobia, anorexia nervosa, anxious insomnia, bipolar disorder, burnout, cyclothymia, generalized anxiety disorder, mood disorder, obsessive-compulsive disorder, post-traumatic stress disorder, panic disorder, psychosis, and schizophrenia.

### Exposure definitions

All registered medications, including OTC products, vaccines, and folic acid, were analysed. Medications were defined as registered pharmaceutical products containing one or more active ingredients available on the Belgian market at the time of data extraction, and were coded according to the WHO Anatomical Therapeutic Chemical (ATC) classification system (preferably 5th level, lower if detail lacked). ATC codes were retrieved automatically from SAM, and manually coded if needed.

Folic acid exposure was defined as use of any product containing ≥0.4 mg, as per the product leaflet. Products with ≥4 mg per tablet were classified as high-dose. Supplements such as vitamin D or iron were classified as medications only if listed in SAM. Free-text entries labelled as “folic acid” or “pregnancy vitamins” with missing dose or product name were assumed to contain ≥0.4 mg, since it is the lowest available dose on the Belgian market. Coding of folic acid exposures was performed manually.

Exposure timing was categorized into four pregnancy periods: the preconception period (defined as six months before the last menstrual period [LMP] until LMP), and the three trimesters of pregnancy (T1: day 0–84; T2: day 85–182; T3: day 183–280). Only exposures with a start day strictly before the delivery date were classified as in-pregnancy.

### Other variables

The following characteristics were extracted from the enrolment questionnaire (Supplementary Table 1): gestational age at enrolment, language of questionnaire completion, maternal age at LMP, ethnic background, civil status, highest educational attainment, employment status in the past year, gross annual household income, gravidity, method of conception, multiple gestation, and the presence of chronic conditions prior to the start of pregnancy.

### Data preparation and analysis

Self-reported exposures were harmonised into a standard analytic dataset using validated R scripts (available via https://gitlab.kuleuven.be/pharmacology/belpreg-drug-utilization-study). Each reported product generated a structured exposure record with start/stop dates relative to LMP. LMP was determined hierarchically: (1) subtracting 280 days from the expected delivery date recorded in the postpartum questionnaire, (2) from ultrasound-or fertility-based due date registered at enrolment, or (3) from self-reported LMP. Potentially implausible entries (e.g. stop date before start date, exposures far beyond delivery, invalid LMP) were corrected where clear entry errors were identified.

The dataset was initially structured at the exposure level (one row per product). For trimester-specific analyses, it was restructured to participant level, generating binary exposure indicators (‘any use’) per period and calculating duration of use. If the exact timing was missing but trimester-level exposure was indicated, the exposure contributed to prevalence but not duration analyses.

Descriptive statistics summarised participant characteristics and prevalence of medication, vaccine, and folic acid use. Concurrent medication use was quantified by counting distinct ATC-coded (lowest level available) products per trimester. Medication prevalence was reported by ATC level 1 categories, with additional analysis of the ten most common ATC level 2 groups. For selected therapeutic classes, prevalence and duration were examined at ATC levels 3–5. Median, interquartile range (IQR), minimum, and maximum durations were reported for T1–T3. For folic acid, initiation timing was categorised into groups based on the earliest start date. Guideline-concordant use (per Belgian recommendations) required initiation ≥28 days before LMP and at least 85 days of use during T1 [16]. Daily exposure sequences were merged to avoid double-counting overlapping or adjacent exposures.

Missing covariate data were retained as explicit “missing” categories. All analyses were performed in R (version 2025.05.0).

## 4. Results

A total of 2,096 participants completed follow-up beyond T1 and were included in the base cohort; 1,767 were followed through T2 and 1,136 through T3 (Table 1). Median gestational age at enrolment was 16 weeks (IQR 10–25). Most were Dutch-speaking (≈91%), aged 30–34 years (≈52%), Caucasian (≈98%), and partnered (≈98%). The majority reported higher education (≈88%), employment (≈97%), and planned pregnancies (≈92%). About 17% conceived through assisted reproductive technology (ART), and ≈47% were primigravida. Self-reported chronic conditions prior to pregnancy were common (40%), most often allergic/hypersensitivity, immune-mediated inflammatory diseases, and thyroid disorders (Supplementary Table 2).

**Table 2.** Prevalence of ATC level 1 medication use before pregnancy and across pregnancy trimesters.

|  | Preconception<br>(N = 2096) | Trimester 1<br>(N = 2096) | Trimester 2<br>(N = 1767) | Trimester 3<br>(N = 1136) |
| --- | --- | --- | --- | --- |
|  | n (%) | n (%) | n (%) | n (%) |
| **Any medication, including vaccines (J07)** | **1681 (80.2%)** | **1798 (85.8%)** | **1625 (92.0%)** | **1078 (94.9%)** |
| **Any medication, excluding vaccines (J07)** | **1635 (78.0%)** | **1756 (83.8%)** | **1575 (89.1%)** | **973 (85.7%)** |
| A – Alimentary tract and metabolism | 312 (14.9%) | 508 (24.2%) | 551 (31.2%) | 431 (37.9%) |
| B – Blood and blood forming organs | 218 (10.4%) | 392 (18.7%) | 525 (29.7%) | 387 (34.1%) |
| C – Cardiovascular system | 71 (3.4%) | 68 (3.2%) | 76 (4.3%) | 104 (9.2%) |
| D – Dermatologicals | 54 (2.6%) | 71 (3.4%) | 81 (4.6%) | 46 (4%) |
| G – Genito urinary system and sex hormones | 568 (27.1%) | 314 (15.0%) | 157 (8.9%) | 83 (7.3%) |
| H – Systemic hormonal preparations, excluding sex hormones and insulins | 217 (10.4%) | 256 (12.2%) | 218 (12.3%) | 158 (13.9%) |
| J – Antiinfectives for systemic use | 271 (12.9%) | 362 (17.3%) | 639 (36.2%) | 818 (72%) |
| L – Antineoplastic and immunomodulating agents | 150 (7.2%) | 110 (5.2%) | 29 (1.6%) | 14 (1.2%) |
| M – Musculo-skeletal system | 206 (9.8%) | 91 (4.3%) | 21 (1.2%) | 1 (0.1%) |
| N – Nervous system | 940 (44.8%) | 894 (42.7%) | 717 (40.6%) | 320 (28.2%) |
| P – Antiparasitic products, insecticides and repellents | 15 (0.7%) | 16 (0.8%) | 11 (0.6%) | 5 (0.4%) |
| R – Respiratory system | 433 (20.7%) | 886 (42.3%) | 788 (44.6%) | 331 (29.1%) |
| S – Sensory organs | 12 (0.6%) | 12 (0.6%) | 21 (1.2%) | 7 (0.6%) |
| V – Various | 8 (0.4%) | 5 (0.2%) | 3 (0.2%) | 1 (0.1%) |

All results are shown as absolute numbers (n) and percentages (%).

Medication use was highly prevalent across all periods. In the six months before conception, 80.2% of participants reported at least one medication, increasing to 85.8% in T1, 92.0% in T2, and 94.9% in T3. Excluding vaccines, prevalence still ranged between 83.8% and 89.1%. The number of distinct products rose with gestational age, with the median increasing from two in the preconception period to three in T3 (Supplementary Figures 1–2). The most frequently used ATC level 1 categories were nervous system, respiratory system, blood and blood-forming organs, alimentary tract and metabolism, and systemic anti-infectives. At ATC level 2, analgesics, vaccines, antihistamines, antianemic preparations, and drugs for acid-related disorders were most common (Tables 3). Chronic therapies such as thyroid medication and antianemics (with high dose folic acid being most common before and during T1, and ferrous fumarate from T2 onward) were generally used for longer durations, while acute medications like analgesics and antibiotics were typically short-term. A complete list of prevalence rates, median durations, and IQRs for all ATC Level 2 medications used before and during pregnancy is provided in Supplementary Table 3.

**Table 3.** Top 10 ATC level 2 medication groups used during pregnancy.

|  | Trimester 1 (N = 2096) |  |  | Trimester 2 (N = 1767) |  |  | Trimester 3 (N = 1136) |  |  |
| --- | --- | --- | --- | --- | --- | --- | --- | --- | --- |
|  | n (%) | Duration (days) |  | n (%) | Duration (days) |  | n (%) | Duration (days) |  |
|  |  | Median | IQR |  | Median | IQR |  | Median | IQR |
| **N02 – Analgesics** | 774 (36.9%) | 3 | 4 | 626 (35.4%) | 3 | 3 | 247 (21.7%) | 3 | 5 |
| **J07 – Vaccines** | 274 (13.1%) | 1 | 0 | 515 (29.1%) | 1 | 0 | 785 (69.1%) | 1 | 0 |
| **R06 – Antihistamines for systemic use** | 755 (36.0%) | 34 | 27 | 620 (35.1%) | 52 | 78 | 222 (19.5%) | 82 | 76.8 |
| **B03 – Antianemic preparations** | 259 (12.4%) | 80.5 | 40 | 303 (17.1%) | 15 | 47 | 271 (23.9%) | 77 | 48 |
| **A02 – Drugs for acid-related disorders** | 182 (8.7%) | 15.5 | 78.8 | 302 (17.1%) | 10 | 50 | 311 (27.4%) | 30 | 79 |
| **H03 – Thyroid therapy** | 214 (10.2%) | 85 | 38 | 202 (11.4%) | 98 | 7.2 | 145 (12.8%) | 94.5 | 14.8 |
| **B01 – Antithrombotic agents** | 146 (7.0%) | 34 | 49 | 263 (14.9%) | 95 | 13 | 156 (13.7%) | 70 | 9 |
| **R01 – Nasal preparations** | 175 (8.3%) | 14 | 80 | 210 (11.9%) | 10 | 40 | 121 (10.7%) | 11 | 59 |
| **G03 – Sex hormones and modulators of the genital system** | 295 (14.1%) | 38 | 54 | 118 (6.7%) | 13.5 | 47.8 | 53 (4.7%) | 51 | 41 |
| **J01 – Antibacterials for systemic use** | 86 (4.1%) | 7.5 | 5 | 165 (9.3%) | 6 | 4 | 90 (7.9%) | 6 | 5 |

All results are shown as absolute numbers (n) and percentages (%); ordered by average rank based on percentage of users in each trimester; IQR: interquartile range.

Table 4 details the prevalence and duration of selected therapeutic classes. Analgesics were widely used, with prevalence peaking at 36.9% in T1 and declining to 21.7% in T3. This was almost entirely attributable to paracetamol, already common preconception (36.5%) and reported by 35.4%, 34.4%, and 21.0% of participants in the T1, T2, and T3, respectively. Median durations of paracetamol remained stable at three days (IQR 3–5). Antidepressants were used by around 5% throughout pregnancy, with long durations consistent with chronic treatment. Antipsychotics, stimulants, and hypnotics or sedatives were rarely used; the latter, when reported, had moderate durations of 24–32 days.

**Table 4.**
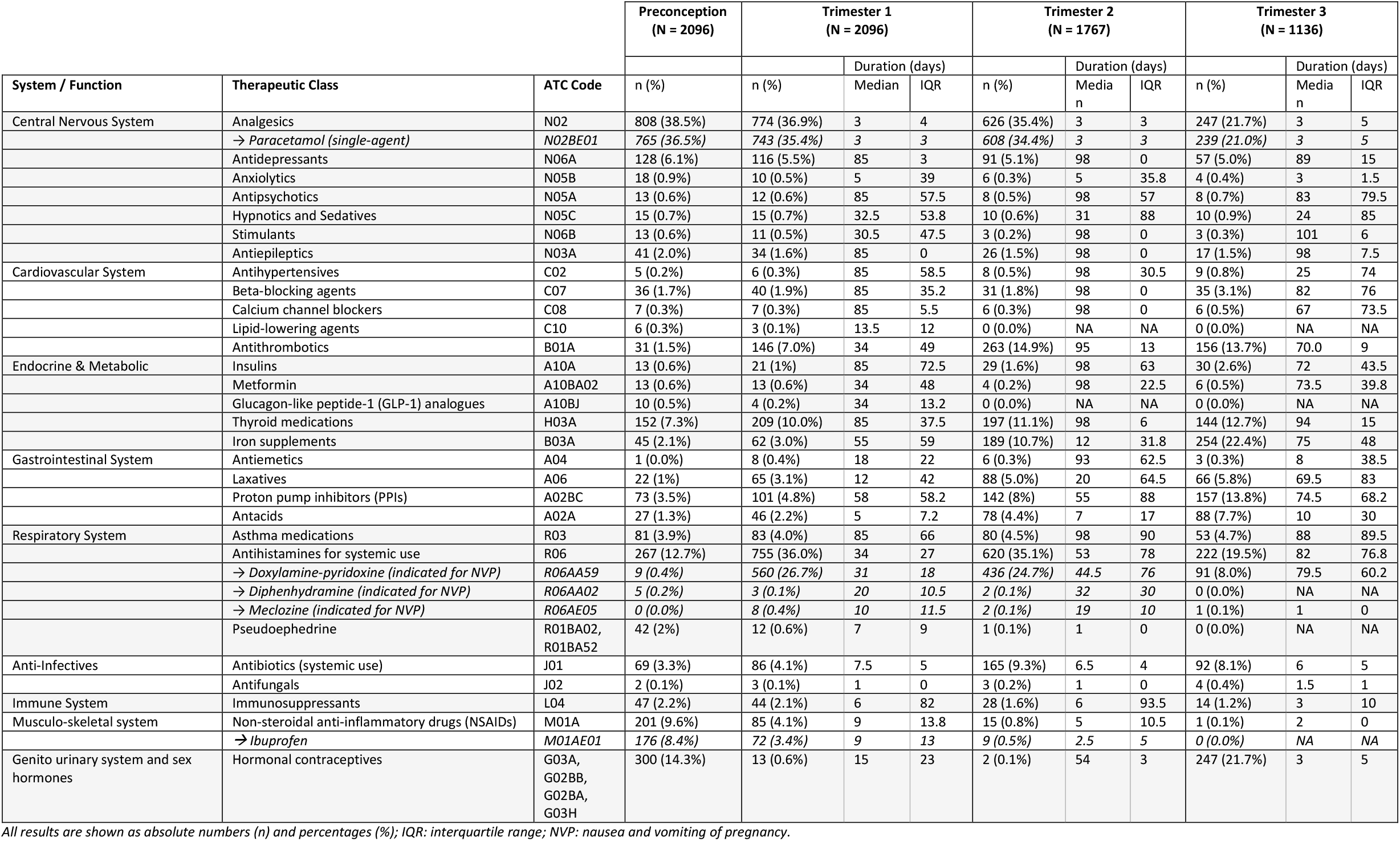
Prevalence and duration of selected therapeutic classes, and individual active substances, before and during pregnancy by trimester.

Cardiovascular medication use increased during pregnancy, particularly the ATC group of antithrombotics, which rose from 7.0% in T1 to 14.9% in T2, largely explained by the registration of low-dose aspirin use. Beta-blocker use was low but increased slightly to 3.1% in T3. In the endocrine/metabolic category, thyroid medication use rose from 7.3% preconceptionally to 10.0% in T1 and 12.7% in T3, with consistently long durations. Iron supplementation increased markedly from 2.1% before conception to 22.4% in T3. Insulin use rose modestly to 2.6% in T3, while oral hypoglycaemics, including metformin (≤0.6% throughout pregnancy), and GLP-1 analogues (0.2% in T1 and absent later in pregnancy), remained rare.

Gastrointestinal medication use became more prevalent with increasing gestational age: by T3, 13.8% of participants reported proton pump inhibitor use, 7.7% used antacids, and 5.8% used laxatives. Antiemetic use was largely driven by the doxylamine-pyridoxine combination, which was reported by 26.7% in T1, 24.4% in T2, and 8.0% in T3, with median duration increasing across trimesters. Diphenhydramine and meclozine were rarely used (≤0.4%). Systemic antibiotics were reported by 9.3% of participants in T2 and 8.1% in T3, with short durations.

Exposure to potentially inappropriate or teratogenic medications was rare. NSAID use declined from 4.1% in T1 to 0.8% in T2 and 0.1% in T3, and predominantly describes ibuprofen use. Only one participant reported ACE inhibitor use in T1 (0.05%, duration of 28 days), no other teratogenic medications were reported during pregnancy (Supplementary List 1). Topiramate, methotrexate, and angiotensin II receptor blockers were only (rarely) recorded before conception.

Vaccine uptake was substantial. Pertussis vaccination was mostly reported in T3 (57.2%, median GA 206 days, ± 29 weeks), with an additional 9.9% in T2. Influenza vaccination occurred across trimesters, most frequently reported in T2 (18.1%), followed by T1 (10.3%) and T3 (13.1%). COVID-19 vaccination followed a similar pattern, with 10.3% in T2, 5.3% in T1, and 5.9% in T3. The maternal RSV vaccine, was reported by 5.3% in T3 (median GA 237 days). Other vaccines were rarely used in pregnancy, usually before conception (Supplementary Table 4).

Folic acid supplementation in pregnancy was nearly universal (Table 5). Regarding the timing of folic acid initiation, 65.0% initiated use ≥28 days before LMP, 4.8% between 14 and 28 days before LMP, and 15.9% after 28 days post-LMP. Guideline-concordant use, defined as timely initiation plus ≥85 days of use in the first trimester, was achieved by 59.9% of participants (of the 1355 participants who initiated supplementation timely, 107 did not meet the adherence threshold of 85 days in T1). High-dose folic acid (≥4 mg) was reported by 9.7% in T1, 6.3% in T2, and 1.0% in T3.

**Table 5.** Folic acid use across pregnancy trimesters and timing of initiation.

|  | Start ≥28 days before LMP (N=2084<sup>1</sup>) | Start between 28 and 14 days before LMP (N=2084<sup>1</sup>) | Start after 28 days post LMP (N=2084<sup>1</sup>) | Used folic acid concordant to guideline in Belgium (N=2084<sup>1</sup>) | Use in trimester 1 (N = 2096) |  |  | Use in trimester 2 (N = 1767) |  |  | Use in trimester 3 (N = 1136) |  |  |
| --- | --- | --- | --- | --- | --- | --- | --- | --- | --- | --- | --- | --- | --- |
|  | n (%) | n (%) | n (%) | n (%) |  | Duration (days) |  |  | Duration (days) |  |  | Duration (days) |  |
|  |  |  |  |  | n (%) | Median | IQR | n (%) | Median | IQR | n (%) | Median | IQR |
| **Minimum 0.4mg of folic acid<sup>2</sup>** | 1355 (65.0%)<sup>3</sup> | 101 (4.8%)<sup>3</sup> | 332 (15.9%)<sup>3</sup> | 1248 (59.9%) | 2081 (99.3%) | 51 | 39 | 1759 (99.5%) | 75 | 55 | 1128 (99.3%) | 56 | 77 |

All results are shown as absolute numbers (n) and percentages (%); IQR: interquartile range.<sup>1</sup> Includes all pregnancies that were followed until the end of the first trimester, with exact start and stop dates for folic acid available (N = 12 had missing start/stop data in the T1 cohort).<sup>2</sup> Less than 4% of all folic acid registrations consisted of free-text entries labelled as “folic acid” or “pregnancy vitamins” without dosage or product name; these were assumed to contain at least 0.4 mg of folic acid, based on the minimum dosage found in such products currently available on the Belgian market. <sup>3</sup>Regarding timing of initiation, the table presents the following clinically relevant categories: ‘start ≥28 days before LMP’, ‘start between 28 and 14 days before LMP’, and ‘start after 28 days post-LMP’. Categories not shown in the table include: ‘start between 13 days before LMP and LMP’ (n = 92, 4.4%), ‘start after LMP but before 28 days post-LMP’ (n = 184, 8.8%), ‘pregnancies with a stop date ≥28 days before LMP’ (n = 5, 0.2%), and ‘pregnancies with no folic acid registrations in T1’ (n = 15, 0.7%).

## 5. Discussion

This study provides the first comprehensive, prospective overview of medication and vaccine use, as well as folic acid intake, among pregnant individuals in Belgium. Medication use was extremely common, reported by 85.8% in T1 and rising to nearly 95% by T3. These results are at the higher end of international estimates [6] and comparable to data from the Netherlands [17]. Administrative data such as the Belgian BeMeP cohort (2010–2016) reported lower prevalence (78.9%) [13], largely because they capture only dispensed, reimbursed products and miss OTCs. By contrast, BELpREG’s prospective self-report design, with four-week intervals, captures both prescription and OTC use and provides granular information on timing and duration.

Beyond the high overall prevalence, the frequent concurrent use of multiple products, with the median number increasing from two in the first trimester to three in later pregnancy, is notable. This reflects evolving pharmacological needs over gestation, including management of iron deficiency, thrombotic risk, gastrointestinal symptoms, constipation, and preeclampsia prevention. Similar increases were noted in the BeMeP cohort, though the median number of dispensed medications was slightly lower [13].

Paracetamol was the most commonly used medication, reported by over one-third of participants in T1 and one-fifth in T3, consistent with international studies showing use by 40–65% of pregnant women at least once in pregnancy [6, 18, 19]. Use of paracetamol in BELpREG was typically short (median 3 days), similar to the MotherToBaby registry, which reported that nearly 60% of women who used paracetamol did so for fewer than 10 days [18]. A notable discrepancy exists between self-reported paracetamol use and prevalence estimates derived from administrative data. These sources typically report much lower usage rates, around 6–8% [20-22], which may reflect a more selective subgroup of users. Given the ongoing debate about potential associations between paracetamol and neurodevelopmental outcomes, safety studies must account for the heterogeneity of exposure patterns [18] and contextual factors such as familial or psychosocial environment [23-25].

Besides paracetamol, NSAIDs are generally used for pain and fever. NSAID use declined with advancing gestation, from 4.1% in T1 to near absence in T3, which is reassuring given the risks of, for example, premature ductus arteriosus closure and renal impairment later in pregnancy. Ibuprofen accounted for nearly all NSAID use. However, Belgian claims data showed slightly higher third-trimester use, probably reflecting peripartum analgesic prescriptions [26]. Other potentially teratogenic medication exposures were almost absent in BELpREG, with only one ACE inhibitor exposure during (early) pregnancy. These findings contrast with higher estimates from administrative data in France [27], the Netherlands [9, 11], and a European survey-based study [10]. However, such findings are strongly influenced by the variety in criteria used to define ‘teratogenicity’, ‘harm’ or ‘risk’. Currently, there is no universally accepted global classification or list of teratogens, which complicates comparisons across studies.

Use of nausea and vomiting in pregnancy (NVP) treatments was common, largely driven by doxylamine-pyridoxine, - which was used by more than one quarter of the cohort. An international database study (2002–2014) reported lower overall prescription antiemetic use, with the highest prevalence in Canada (17.7%), followed by the United States (14.0%) and the United Kingdom (5.0%) [28]. In the U.S. National Birth Defects Prevention Study, 70% of women reported NVP in the first trimester, of whom 12.2% used treatment [29]. By contrast, doxylamine-pyridoxine use in Belgium, where it is also prescription-only, was considerably higher (26.7% and 24.2% in T1 and T2) and unexpectedly persisted into T3 (8%), despite the typical decline of NVP.

In this cohort, maternal vaccine uptake was substantial, particularly for pertussis, though not universal despite guideline recommendations. Combining all trimesters, 67.2% of pregnant women received a pertussis vaccine (57.2% in T3). Coverage was comparable to earlier Belgian data in a smaller cohort (69.3%, N=481, 2021) [30], and higher than estimates from the U.S. (55.1%, 2017–2021) [31] and the United Kingdom (58.6%, 2023–2024) [32]. Influenza vaccination, assessed across flu seasons between November 2022 and July 2025, was registered in 41.5% of pregnancies in BELpREG. This was similar to earlier Belgian data (47.2%, N=481, 2021) [30] and exceeded U.S. estimates (33%, 2017–2021) [31]. COVID-19 vaccination was less frequent (21.5%) [33], reflecting the post-pandemic period of data collection (2022–2025), while maternal RSV vaccination was rare, consistent with its recent market introduction and lack of reimbursement. Persistent gaps in maternal vaccination coverage in Belgium highlight the need for sustained public health attention and multilevel strategies to improve uptake [34, 35].

Folic acid supplementation was almost universal, yet only 59.9% achieved guideline-concordant use (≥28 days preconception plus ≥85 days in T1), and 65.0% initiated timely supplementation. Compared with international studies, adherence in BELpREG appears somewhat higher: in Ireland and Norway, only 16–28% met recommendations [35,36], while in the Dutch ‘Moeders van Morgen’ cohort 55% did so (using the same definition of timely start, but requiring use until ≥8 weeks after conception, compared to ≥85 days in T1 in this study) [37]. Earlier Belgian findings (56%) align closely with our results. Among ART pregnancies, adherence was notably higher, with >74% meeting guideline criteria (Supplementary Table 5), which may partly explain the relatively favourable overall prevalence. High-dose folic acid (≥4 mg) was reported by ∼10%, though this likely reflects some misclassification (the 4 mg product was listed first in the selector during the first period of data collection). While Belgium seems to perform comparatively well in early folic acid uptake, gaps remain in ensuring full adherence to the recommended timing and duration, and, in the absence of food fortification, folic acid tablets as supplements are required for all pregnancies.

The BELpREG registry offers extensive opportunities for future research. The registry’s richness supports future subgroup analyses, for example by maternal characteristics (e.g., BMI) and health behaviours (e.g., vaccine acceptance, folic acid adherence, polypharmacy). Triangulation with administrative data could be important for validation and, in future research, could further strengthen drug utilisation analyses linking additional maternal characteristics. Future work could also distinguish between medications continued from pre-existing chronic therapy and those newly initiated during pregnancy, as well as examine patterns of ad hoc medication use from preconception into pregnancy. Further detailing the use of psychotropics – for example, linking antidepressant use (∼5% in this cohort) with validated patient reported measures of maternal mental health – would provide valuable insights into how medication use reflects underlying disease burden. While this study focused on utilisation, linking detailed exposure data with outcomes will enable medication safety studies across a wide range of products.

### Strengths and limitations

The BELpREG registry provides comprehensive, detailed data on medication use in pregnancy in Belgium, covering prescription drugs, OTC products, and supplements. Data on self-reported use have shown to align well with prescription records for chronic treatments [36-38], and offer added value by capturing actual use, OTC products, and timing of exposure [38, 39]. The four-weekly questionnaire design in BELpREG minimized recall bias, while date fields allowed precise exposure periods. Participant-driven reporting, independent of healthcare providers, may have reduced socially desirable responses. The trimester-specific nested cohort design, independent of pregnancy outcomes, further limited selection bias.

Limitations include reliance on self-reported data, which, despite the design features and applied corrective measures, may still be subject to recall bias and potential misclassification. Voluntary participation likely introduces selection bias and the cohort is demographically skewed toward Dutch-speaking, highly educated, employed women, limiting generalizability. Compared with national data, BELpREG includes more participants aged 30–34 years, fewer aged 18– 24, and a higher proportion with high education, income, and Caucasian ethnicity [40-42]. ART pregnancies were also overrepresented (17.7% vs. 9.6% nationally) [43]. In addition, only pregnancies followed until the end of T1 were included in the base cohort, so early pregnancy losses and T1 loss to follow-up were not captured. French-speaking participants were underrepresented (7.6%), as registry access in French (and English) began only in 2024. Finally, underreporting of hospital-administered and fertility-related medications is possible due to recall difficulties by participants.

## 6. Conclusions

In this first nationwide prospective study of medication use during pregnancy in Belgium, exposure was nearly universal, ranging from 85.8% to 95% across trimesters. Patterns reflected both chronic treatments and pregnancy-related needs, with particularly high use of paracetamol (≈35%) and doxylamine-pyridoxine (≈27%). Folic acid supplementation was almost universal, although only six in ten participants met the national criteria for guideline-concordant use. Vaccine uptake, especially for pertussis, was substantial but incomplete (67.2%). Reassuringly, exposure to potentially teratogenic or inappropriate medications was rare.

## Author Contributions Statement

**Laure Sillis**: Conceptualization, Data curation, Formal analysis, Funding acquisition, Investigation, Methodology, Project administration, Software, Validation, Visualization, Writing – original draft; **Sien Lenie**: Investigation, Formal analysis, Validation, Writing – review and editing; **Emily Jacobs**: Formal analysis, Writing – review and editing; **Karel Allegaert**: Conceptualization, Writing – review and editing; **Annick Bogaerts**: Conceptualization, Writing – review and editing; **Maarten De Vos**: Conceptualization, Writing – review and editing; **Titia Hompes**: Conceptualization, Writing – review and editing; **Anne Smits**: Conceptualization, Writing – review and editing; **Kristel Van Calsteren**: Conceptualization, Writing – review and editing; **Jan Y Verbakel**: Conceptualization, Writing – review and editing, Supervision; **Veerle Foulon**: Conceptualization, Funding acquisition, Methodology, Resources, Supervision, Writing – review and editing; **Michael Ceulemans**: Conceptualization, Funding acquisition, Investigation, Methodology, Project administration, Resources, Supervision, Validation, Writing – review and editing.

## AI Use Statement

Artificial intelligence (AI) supported tools (Copilot) were employed for writing tasks, including refining language, structuring sections, and assisting in the development and correction of scripts used for data processing and analysis. These scripts were based on ELT code written by Laure Sillis. All scientific content, interpretations, and conclusions were developed by the authors. AI was not used to generate original research findings or perform autonomous data analysis.

## Data availability statement

The data used for this analysis were collected under participants’ informed consent for uses as directed by the study principal investigator and are not available for public access due to ethical and privacy reasons. Requests for use of the data should be directed to the corresponding author.

## Ethical approval and informed consent

Ethical approval for this study was obtained from the Ethics Committee Research UZ/KU Leuven (S66464). All BELpREG participants provided electronic informed consent.

## Additional contributions

The authors express their gratitude to the BELpREG participants who provided data for this study and to healthcare professionals for informing and motivating pregnant individuals about participation in BELpREG. The authors also appreciate the technical support from Jared Houghtaling and Digile, and the partnership with Medipim upgrading the quality of the BELpREG questionnaires and data collection.

## Disclaimer

The terms “individuals”, “participants”, “women,” “woman,” “mother” and “maternal” are used in this manuscript to refer to all those who are pregnant and give birth. The authors acknowledge that not all people who are pregnant and give birth will identify themselves as women.

