## Supplementary Material for "Medication, Vaccine, and Folic Acid Use Among Pregnant Women in Belgium: Insights from the BELpREG Cohort"

**Content Supplementary Material**

***Supplementary Table 1.*** *Overview of BELpREG variables used in this study.*

| **Category 1: Sociodemographics** | | | | | | | |
|---|---|---|---|---|---|---|---|
| **Variable** | **Definition** | **Values** | **Field type** | **Source** | **Survey Instruments** | | |
| | | | | | **EM** | **FU** | **PP 1** |
| **BELpREG record id** | Unique numeric code | Integer (Automatically assigned) | Text box | Automatically assigned | x | x | x |
| **Language** | Language of completing the questionnaires | Dutch; French; English | Text box | Reported | x | | |
| **Date of birth** | Date of birth participant | Date (DD-MM-YYYY) | Text box | Reported | x | | |
| **Age** | Age of the participant at first day of the LMP[1] | Integer | Calculated field | *Derived* | | | |
| **Partner** | Currently having a partner | Y/N | Radio buttons | Reported | x | | |
| **Ethnicity participant** | Ethnicity of the participant | European; Maghreb; Rest of Africa; Near/Middle East; Pacific/Far East; Rest of Asia; North America; Central and South America; I don't know / I prefer not to say | Checkboxes, Multiple choice | Reported | x | | |
| **Highest educational level of participant** | Highest educational degree obtained by participant | Primary education; Lower secondary education (certificate 2nd degree); Higher secondary education; Higher vocational education (HBO5); Professional Bachelor / Higher education short type (non-university); Academic Bachelor; Master; PhD; Other (specify) | Radio buttons | Reported | x | | |
| **Professional activity participant** | The participant being professional active in the past year | Y/N | Radio buttons | Reported | x | | |
| **Gross Annual Family Income** | Gross Annual Family Income | <15 000 euro; 15 000 euro - <30 000 euro; 30 000 euro - <45 000 euro; 45 000 euro - <65 000 euro; >=65 000 euro; I don't know / I'd rather not tell | Radio buttons | Reported | x | | |

Abbreviations : EM = Pregnancy Enrolment Questionnaire; FU = Pregnancy Follow-up Questionnaire; PP 1 = First Postpartum Questionnaire
Notes: [1] In case the last menstrual period (LMP) is not directly registered by participants, it is derived from estimated date of delivery (EDD)

| **Category 2: Current pregnancy and health status** | | | | | | | |
|---|---|---|---|---|---|---|---|
| **Variable** | **Definition** | **Values** | **Field type** | **Source** | **Survey Instruments** | | |
| | | | | | **EM** | **FU** | **PP 1** |
| **Estimated date of delivery (EDD)** | Estimated date of delivery | Date (D-M-Y) and 'I don't know yet' checkbox | Text box and Checkbox | Reported | x | | |

| | | | | | | | |
|---|---|---|---|---|---|---|---|
| **Source of the estimated date of delivery** | Method used to define the estimated date of delivery | Ultrasound; Calculated based on the first day of the last period; Calculated based on the date of conception (in case of fertility treatment); Other (Specify) | Radio buttons with embedded Text box | Reported | x | | |
| **Last Menstrual Period (LMP)** | Date of the first day of the last menstrual period prior to conception | Date (D-M-Y) | Text box | Reported | x | | |
| **Onset of pregnancy** | Onset of the current pregnancy | Spontaneously; After fertility treatment with hormonal stimulation; After fertility treatment without hormonal stimulation | Radio buttons | Reported | x | | |
| **Singleton or multiple pregnancy** | Singleton or multiple pregnancy | Singleton; Multiples; I don't know yet | Radio buttons | Reported | x | | |
| **Planned pregnancy** | Planned pregnancy | Y/N | Radio buttons | Reported | x | | |
| **Chronic conditions** | Chronic conditions (i.e., conditions existing before the start of the pregnancy) | Text - selection from drop-down list[2] or free text - multiple answers possible | Drop-down list linked to database, Autocomplete | Reported | x | | |
| **Pregnancy complications** | Pregnancy complications (since previous questionnaire) | No complication(s) or disease(s); Severe pregnancy vomiting (hyperemesis gravidarum); Diabetes diagnosed after 20 weeks ('gestational diabetes'); Increased blood pressure after 20 weeks ('Gestational Hypertension'); Pre-eclampsia diagnosed before/during week 34; Pre-eclampsia diagnosed after week 34; HELLP syndrome; Seizures; Growth retardation in the unborn child; Blood clot (thrombosis); Placenta praevia; Placenta abruptio; (severe) bleeding (specify); Depression during pregnancy; Anxiety during pregnancy; Psychosis; Exacerbation of my chronic condition (specify which condition this concerns); Pregnancy cholestasis; Too much amniotic fluid; Too little amniotic fluid; Threatening premature birth; Premature ruptured of membranes (PROM); Premature labour (< 37 weeks); Infection (specify); Other (specify) | Checkboxes with embedded text fields, Multiple choice | Reported | x | x | x |

Abbreviations : EM = Pregnancy Enrolment Questionnaire; FU = Pregnancy Follow-up Questionnaire; PP 1 = First Postpartum Questionnaire; EDD = Estimated Date of Delivery; LMP = Last Menstrual Period; ICD-11 = International Classification of Diseases 11th Revision; MedDRA = Medical Dictionary for Regulatory Activities; HELLP = Haemolysis, Elevated Liver enzymes and Low Platelets.

Notes: [1] If the estimated date of delivery (EDD) is not known, the calculation of gestational age is based on the last menstrual period (LMP); [2] Structured text field with autocomplete function, linked to a self-developed list of the most frequently reported disorders linked to their respective ICD-11 and MedDRA classification.

| **Category 3: Maternal-obstetric history** | | | | | | | |
|---|---|---|---|---|---|---|---|
| **Variable** | **Definition** | **Values** | **Field type** | **Source** | **Survey Instruments** | | |
| | | | | | **EM** | **FU** | **PP 1** |
| **Previous pregnancy** | Previously having been pregnant | Y/N | Radio buttons | Reported | x | | |
| **Gravidity** | Gravidity | Primigravida; Multigravida | Calculated field | Derived | | | |

Abbreviations : EM = Pregnancy Enrolment Questionnaire; FU = Pregnancy Follow-up Questionnaire; PP 1 = First Postpartum Questionnaire

| **Category 4: Use of medicines, folic acid / pregnancy vitamins and other health products** | | | | | | | |
|---|---|---|---|---|---|---|---|
| **Variable** | **Definition** | **Values** | **Field type** | **Source** | **Survey Instruments** | | |
| | | | | | **EM** | **FU** | **PP 1** |
| **Medication use during pregnancy** - The following questions are repeated for each reported medicine | | | | | | | |
| Name of the medicine | Name of the medicine used since the start of the pregnancy (i.e., in the enrolment questionnaire or in the postpartum 1 questionnaire) | Text - selection from drop-down list[1] or free text | Drop-down list linked to database, Autocomplete | Reported | x | x | x |
| Modality of medicine use | How was/has the medicine been used | a) Daily and still (e.g., thyroid hormone);<br>b) Occasionally or as needed (e.g., a painkiller taken occasionally, or an injection that is regularly given);<br>c) During a certain period and not anymore (e.g., an antibiotic that was used for 5 days) | Radio buttons | Reported | x | x | x |
| ***Medication use during pregnancy – modality 'a) Daily and still'*** | | | | | | | |
| Start of use | Timing of medication use initiation | Before this pregnancy; During this pregnancy | Radio buttons | Reported | x | | |
| Start of use during pregnancy – date | Start date of medication use during this pregnancy | Date (DD-MM-YYYY) | Text box | Reported | x | x | x |
| Continuation of use | Continuation of a previously registered medicine under the modality 'Daily and still'[3]. | Yes; I no longer use this medicine (if so, when did you stop); The use of this medicine has changed (if so, date of change is questioned) | Radio buttons | Reported | | x | x |
| ***Medication use during pregnancy - modality 'b) Occasionally or as needed'*** | | | | | | | |
| Number of days when medicine was used | Number of days when the medicine was used since the start of the pregnancy (i.e., in the | - During the first trimester (from week 1 to the end of week 12): integer | Text box (3) | Reported | x | x | x |

| | | | | | | | |
|---|---|---|---|---|---|---|---|
| | enrolment questionnaire) or since completing the previous questionnaire (i.e., in a follow-up questionnaire or in the postpartum 1 questionnaire) | - During the second trimester (from week 13 to the end of week 26): integer<br>- During the third trimester (from week 27 until the end of the pregnancy): integer | | | | | |
| Start of use during pregnancy - date | Start date of medication use during this pregnancy | Date (DD-MM-YYYY) | Text box | Reported | x | x | x |
| Use in 6 months before start pregnancy | Use of the medicine in the 6 months before the start of the pregnancy | Y/N | Radio buttons | Reported | x | | |
| ***Medication use during pregnancy - modality 'c) During a certain period and not anymore'*** | | | | | | | |
| Start of use | Timing of medication use initiation | Before this pregnancy; During this pregnancy | Radio buttons | Reported | x | x | x |
| Start of use during pregnancy - date | Start date of medication use during this pregnancy | Date (DD-MM-YYYY) | Text box | Reported | x | x | x |
| Stop of use | Stop date of medication use | Date (DD-MM-YYYY) | Text box | Reported | x | x | x |
| **Use of folic acid and pregnancy vitamins during pregnancy** - The following questions are repeated for each reported folic acid product and pregnancy vitamin | | | | | | | |
| **Name of the folic acid product or pregnancy vitamin** | Name of the folic acid product or pregnancy vitamin used currently (i.e., in the enrolment questionnaire) or since completing the previous questionnaire (i.e., in a follow-up questionnaire or in the postpartum 1 questionnaire) | Text – selection from drop-down list[1] or free text | Drop-down list linked to database, Autocomplete | Reported | x | x | x |
| Start of use | Start date of using the folic acid product or pregnancy vitamin | Date (DD-MM-YYYY) | Text box | Reported | x | x | x |
| Stop of use | Stop date of using the folic acid product or pregnancy vitamin | Date (DD-MM-YYYY) | Text box | Reported | | x | x |
| Continuation of use | Continuation of a previously registered folic acid product or pregnancy vitamin[4] | Yes; I no longer use this product (if so, when did you stop); The use of this product has changed (if so, date of change is questioned) | Radio buttons | Reported | | x | x |
| **Name of a folic acid product or pregnancy vitamin used earlier in pregnancy** | Name of a folic acid product or pregnancy vitamin used earlier in this pregnancy | Text - selection from drop-down list[1] or free text | Drop-down list linked to database, Autocomplete | Reported | x | | |
| Start of use | Start date of using the folic acid product or pregnancy vitamin | Date (DD-MM-YYYY) | Text box | Reported | x | | |
| Stop of use | Stop date of using the folic acid product or pregnancy vitamin | Date (DD-MM-YYYY) | Text box | Reported | x | | |
| **Health products' use during pregnancy** - The following questions are repeated for each reported health product | | | | | | | |

| | | | | | | | |
|---|---|---|---|---|---|---|---|
| **Name of the health product** | Name of the health product used since the start of the pregnancy (i.e., in the enrolment questionnaire) or since completing the previous questionnaire (i.e., in a follow-up questionnaire or in the postpartum 1 questionnaire) | Text - selection from drop-down list[1] or free text | Drop-down list linked to database, Autocomplete | Reported | x | x | x |
| **Modality of health product use** | How was/has the health product been used | a) Daily and still (e.g., thyroid hormone);<br>b) Occasionally or as needed (e.g., a painkiller taken occasionally, or an injection that is given regularly);<br>c) During a certain period and not anymore (e.g., an antibiotic that was used for 5 days) | Radio buttons | Reported | x | x | x |
| ***Health products' use during pregnancy - modality 'a) Daily and still'*** | | | | | | | |
| Start of use | Timing of the initiation of the use of the health product | Before this pregnancy; During this pregnancy | Radio buttons | Reported | x | | |
| Start of use during pregnancy - date | Start date of using the health product during this pregnancy | Date (DD-MM-YYYY) | Text box | Reported | x | x | x |
| Continuation of use | Continuation of a previously registered health product under the modality 'Daily and still'[2]. | Yes; I no longer use this product (if so, when did you stop); The use of this product has changed (if so, date of change is questioned) | Radio buttons | Reported | | x | x |
| ***Health products' use during pregnancy - modality 'b) Occasionally or as needed'*** | | | | | | | |
| Number of days when heath product used | Number of days when the health product was used since the start of the pregnancy (i.e., in the enrolment questionnaire) or since completing the previous questionnaire (i.e., in a follow-up questionnaire or in the postpartum 1 questionnaire) | - During the first trimester (from week 1 to the end of week 12): integer<br>- During the second trimester (from week 13 to the end of week 26): integer<br>- During the third trimester (from week 27 until the end of the pregnancy): integer | Text box (3) | Reported | x | x | x |
| Start of use during pregnancy - date | Start date of the use of the health product during this pregnancy | Date (DD-MM-YYYY) | Text box | Reported | x | x | x |
| Use in 6 months before start pregnancy | Use of the health product in the 6 months before the start of the pregnancy | Y/N | Radio buttons | Reported | x | | |
| ***Health products' use during pregnancy - modality 'c) During a certain period and not anymore'*** | | | | | | | |
| Start of use | Timing of the initiation of the use of the health product | Before this pregnancy; During this pregnancy | Radio buttons | Reported | x | x | x |
| Start of use during pregnancy - date | Start date of the use of the health product during this pregnancy | Date (DD-MM-YYYY) | Text box | Reported | x | x | x |
| Stop of use | Stop date of the use of the health product | Date (DD-MM-YYYY) | Text box | Reported | x | x | x |

| | | | | | | | |
|---|---|---|---|---|---|---|---|
| **Other medicines and health products used in the 6 months before the start of the pregnancy** | | | | | | | |
| **Name of the medicine or health product** | Name of the medicine or health product used in the 6 months before the start of the pregnancy | Text - selection from drop-down list[1] or free text | Drop-down list linked to database, Autocomplete | Reported | x | | |
| Stop of use | Stop date of the use of the medicine or health product (estimation) | Date (DD-MM-YYYY) | Text box | Reported | x | | |
| **Vaccine use during pregnancy** | | | | | | | |
| Type of the vaccine | Type of the vaccine used since the start of the pregnancy (i.e., in the enrolment questionnaire) or since completing the previous questionnaire (i.e., in a follow-up questionnaire or in the postpartum 1 questionnaire) | Pertussis vaccine; Influenza vaccine; COVID vaccine; RSV vaccine; Travel-related vaccine; Another vaccine | Checkbox, , Multiple choice | Reported | x | x | x |
| Name of the vaccine | Name of the vaccine used since the start of the pregnancy (i.e., in the enrolment questionnaire) or since completing the previous questionnaire (i.e., in a follow-up questionnaire or in the postpartum 1 questionnaire) | Text - selection from drop-down list[1], only showing names of the selected type of vaccine, or free text - Repeated for each selected vaccine | Drop-down list linked to database, Autocomplete | | x | x | x |
| Date of the vaccination | Date of the vaccination | Date (DD-MM-YYYY) - Repeated for each selected vaccine | Text box | Reported | x | x | x |

Abbreviations : EM = Pregnancy Enrolment Questionnaire; FU = Pregnancy Follow-up Questionnaire; PP 1 = First Postpartum Questionnaire; ATC = Anatomical Therapeutic Chemical; CNK = National Code Number; VMP = Virtual Medicinal Product; AMP = Actual Medicinal Product; NMP = Non Medicinal Product; ICD-11 = International Classification of Diseases 11th Revision; MedDRA = Medical Dictionary for Regulatory Activities. Notes: [1]Structured text field with autocomplete function, including pictures of the products, linked to a database of available medicines and medicinal products in Belgium. Additional information of the product is derived from the linked database, including ATC code, CNK, VMP, VMP group, Substance, Substance strength, Route of drug administration, Pharmaceutical form, Medication (AMP) / non-medication status; [2]If a medicine or health product was registered as 'Daily and still' in the previous questionnaire, this medicine will be shown with the reported details of use in the next questionnaire. This question is always shown first, after which other products can be registered; [3]Structured text field with autocomplete function, linked to a self-developed list of the most frequent reported indications / conditions that are linked to their respective ICD-11 and MedDRA classification; [4]If a folic acid product or pregnancy vitamin was registered in the previous questionnaire, this medicine will be shown with the reported details of use in the next questionnaire. This question is always shown first, after which other products can be registered.

***Supplementary Table 2.*** *Chronic conditions existing prior to pregnancy onset as reported by BELpREG participants.*

| | Follow-up period until the end of trimester 1 (N = 2096) | | Follow-up period until the end of trimester 2 (N = 1767) | | Follow-up period until the end of trimester 3 (N = 1136) | |
|---|---|---|---|---|---|---|
| | n | % | n | % | n | % |
| **Any Chronic Condition** | **841** | **40.1** | **707** | **40.0** | **448** | **39.4** |
| Allergic & Hypersensitivity Conditions | 222 | 10.6 | 195 | 11.0 | 125 | 11.0 |
| Immune-mediated Inflammatory diseases | 184 | 8.8 | 150 | 8.5 | 94 | 8.3 |
| Thyroid Disorders | 130 | 6.2 | 113 | 6.4 | 76 | 6.7 |
| Gastrointestinal Diseases | 120 | 5.7 | 101 | 5.7 | 65 | 5.7 |
| Reproductive & Gynaecological Disorders | 96 | 4.6 | 69 | 3.9 | 50 | 4.4 |
| Asthma | 92 | 4.4 | 82 | 4.6 | 55 | 4.8 |
| Headache & Migraine Syndromes | 89 | 4.3 | 78 | 4.4 | 54 | 4.8 |
| Musculoskeletal & Connective Tissue Disorders | 64 | 3.1 | 52 | 2.9 | 34 | 3.0 |
| Other Mental Health Disorders[1] | 60 | 2.9 | 45 | 2.6 | 33 | 2.9 |
| Cardiovascular & Circulatory Diseases | 56 | 2.7 | 43 | 2.4 | 28 | 2.5 |
| Diabetes & Insulin Resistance | 40 | 1.9 | 38 | 2.2 | 16 | 1.4 |
| Coagulation & Blood Disorders | 33 | 1.6 | 29 | 1.6 | 19 | 1.7 |
| Other Respiratory & Pulmonary Disorders[2] | 33 | 1.6 | 30 | 1.7 | 18 | 1.6 |
| Depression and Depressive Disorders | 26 | 1.2 | 24 | 1.4 | 13 | 1.1 |
| Neuropathies & Nerve Disorders | 25 | 1.2 | 21 | 1.2 | 12 | 1.1 |
| Epilepsy | 24 | 1.2 | 22 | 1.3 | 16 | 1.4 |
| Neurodevelopmental Disorders | 23 | 1.1 | 15 | 0.9 | 10 | 0.9 |
| Genetic & Chromosomal Syndromes | 18 | 0.9 | 16 | 0.9 | 10 | 0.9 |
| Metabolic & Nutritional Disorders | 15 | 0.7 | 13 | 0.7 | 9 | 0.9 |
| Kidney & Urinary Disorders | 12 | 0.6 | 10 | 0.6 | 4 | 0.4 |
| Oncological Disease (Benign & Malignant) | 10 | 0.5 | 9 | 0.5 | 7 | 0.6 |
| Infectious Diseases | 8 | 0.4 | 7 | 0.4 | 2 | 0.2 |
| Other Endocrine Disorders[3] | 7 | 0.3 | 7 | 0.4 | 4 | 0.4 |
| Dermatological Conditions | 7 | 0.3 | 6 | 0.3 | 2 | 0.2 |

*All results are shown as absolute numbers (n) and percentages (%). Chronic conditions existing prior to pregnancy onset were self-reported (and known) by BELpREG participants during the enrolment questionnaire. They could select conditions from a drop-down menu, utilize a structured text field with an autocomplete function linked to a self-developed list of frequently reported disorders mapped to their corresponding ICD-11 codes, or enter conditions as free text. Multiple responses were possible, the field could be left blank in case of no conditions. [1]Including addiction, agoraphobia, anorexia nervosa, anxious insomnia, bipolar disorder, burnout, cyclothymia, generalized anxiety disorder, mood disorder, obsessive-compulsive disorder, post-traumatic stress disorder, panic disorder, psychosis, and schizophrenia; [2]Including allergic bronchopulmonary aspergillosis, atypical pulmonary hypersensitivity, bronchiectasis, chronic rhinosinusitis, chronic sinusitis, chronic hyperventilation, chronic upper respiratory inflammation, hyperreactive airways, and post-covid lung conditions; [3]Including cortisol deficiency, diabetes insipidus, growth hormone deficiency, hypoparathyroidism, lymphocytic pituitarism, pituitary cyst, and prolactinoma.*

***Supplementary Figure 1.*** *Distribution of the number of distinct products (most specific ATC level) per individual, by trimester cohort.*

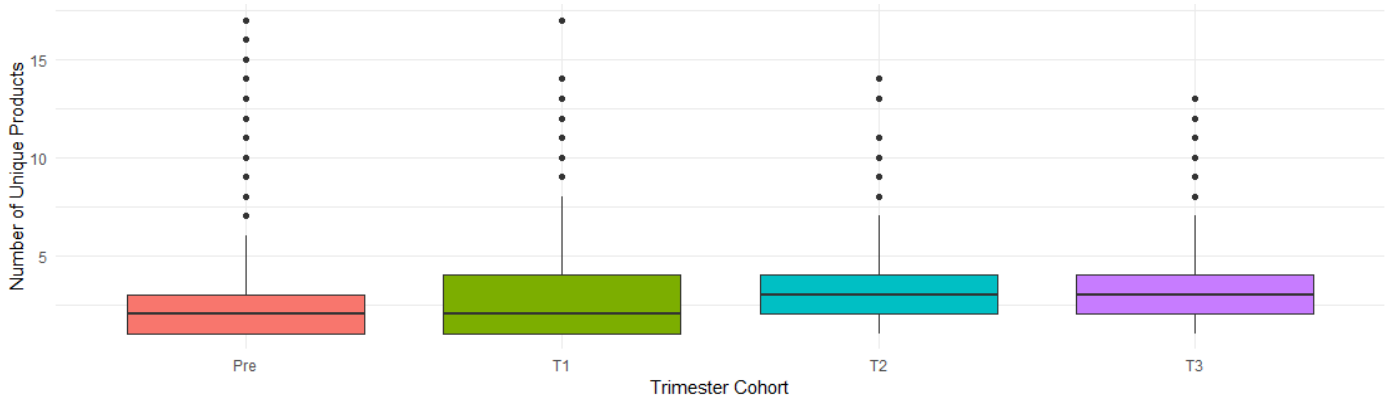

*Pre: preconception (N = 2096); T1 : trimester 1 (N = 2096) ; T2 : trimester 2 (N = 1767); T3: trimester 3 (N = 1136).*

***Supplementary Figure 2.*** *Histogram of the number of distinct medications (lowest available ATC level) per individual, by trimester cohort.*

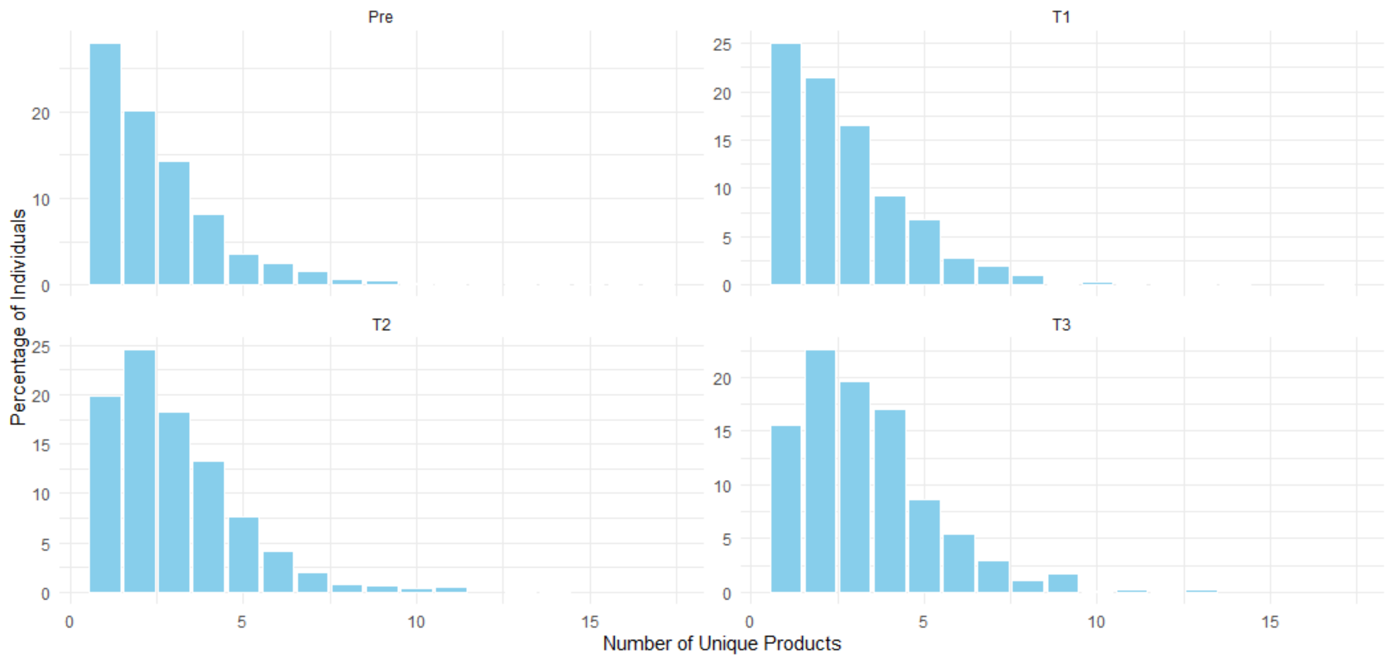

*Pre: preconception (N = 2096); T1 : trimester 1 (N = 2096) ; T2 : trimester 2 (N = 1767); T3: trimester 3 (N = 1136).*

***Supplementary Table 3.*** *Prevalence of ATC level 2 medication use before pregnancy and across pregnancy trimesters.*

| | Preconception (N = 2096) | Trimester 1 (N = 2096) | | | Trimester 2 (N = 1767) | | | Trimester 3 (N = 1136) | | |
|---|---|---|---|---|---|---|---|---|---|---|
| | | | Duration (days) | | | Duration (days) | | | Duration (days) | |
| | n (%) | n (%) | Median | IQR | n (%) | Median | IQR | n (%) | Median | IQR |
| A01 - Stomatological preparations | 1 (0.0%) | 0 (0.0%) | NA | NA | 4 (0.2%) | 7 | 2.5 | 0 (0.0%) | NA | NA |
| A02 - Drugs for acid-related disorders | 129 (6.2%) | 182 (8.7%) | 15.5 | 78.8 | 302 (17.1%) | 10 | 50 | 311 (27.4%) | 30 | 79 |
| A03 - Drugs for functional gastrointestinal disorders | 61 (2.9%) | 166 (7.9%) | 14 | 25 | 94 (5.3%) | 10 | 26.5 | 36 (3.2%) | 8 | 49.5 |
| A04 - Antiemetics and antinauseants | 1 (0.0%) | 8 (0.4%) | 18 | 22 | 6 (0.3%) | 93 | 62.5 | 3 (0.3%) | 8 | 38.5 |
| A05 - Bile and liver therapy | 1 (0.0%) | 1 (0.0%) | 85 | 0 | 0 (0.0%) | NA | NA | 5 (0.4%) | 7 | 11 |
| A06 - Drugs for constipation | 22 (1%) | 65 (3.1%) | 12 | 42 | 88 (5.0%) | 20 | 64.5 | 66 (5.8%) | 69.5 | 83 |
| A07 - Antidiarrheals, intestinal anti-inflammatory/anti-infective agents | 34 (1.6%) | 35 (1.7%) | 15 | 80 | 33 (1.9%) | 9 | 96 | 22 (1.9%) | 25 | 83.8 |
| A09 - Digestives, including enzymes | 2 (0.1%) | 2 (0.1%) | 85 | 0 | 2 (0.1%) | 98 | 0 | 0 (0.0%) | NA | NA |
| A10 - Drugs used in diabetes | 33 (1.6%) | 33 (1.6%) | 70 | 62 | 32 (1.8%) | 98 | 63 | 33 (2.9%) | 72 | 44.5 |
| A11 - Vitamins | 66 (3.1%) | 110 (5.2%) | 12.5 | 46.5 | 88 (5.0%) | 27 | 95 | 44 (3.9%) | 61.5 | 88.2 |
| A12 - Mineral supplements | 4 (0.2%) | 7 (0.3%) | 28 | 59 | 9 (0.5%) | 22.5 | 22 | 3 (0.3%) | 37 | 34 |
| A16 - Other alimentary tract and metabolism products | 1 (0.0%) | 1 (0.0%) | 85 | 0 | 1 (0.1%) | 98 | 0 | 1 (0.1%) | 93 | 0 |
| B01 - Antithrombotic agents | 31 (1.5%) | 146 (7%) | 34 | 49 | 263 (14.9%) | 95 | 13 | 156 (13.7%) | 70 | 9 |
| B02 - Antihemorrhagics | 0 (0.0%) | 2 (0.1%) | 5 | 2 | 1 (0.1%) | 4 | 0 | 0 (0.0%) | NA | NA |
| B03 - Antianemic preparations | 190 (9.1%) | 259 (12.4%) | 80.5 | 40 | 303 (17.1%) | 15 | 47 | 271 (23.9%) | 77 | 48 |
| B05 - Blood substitutes and perfusion solutions | 0 (0.0%) | 0 (0.0%) | NA | NA | 1 (0.1%) | NA | NA | 0 (0.0%) | NA | NA |
| C01 - Cardiac therapy | 1 (0.0%) | 1 (0.0%) | 47 | 0 | 0 (0.0%) | NA | NA | 0 (0.0%) | NA | NA |
| C02 - Antihypertensives | 5 (0.2%) | 6 (0.3%) | 85 | 58.5 | 8 (0.5%) | 98 | 30.5 | 9 (0.8%) | 25 | 74 |
| C03 - Diuretics | 3 (0.1%) | 2 (0.1%) | 5 | 0.5 | 0 (0.0%) | NA | NA | 0 (0.0%) | NA | NA |
| C05 - Vasoprotectives | 12 (0.6%) | 14 (0.7%) | 27 | 75 | 38 (2.2%) | 23 | 61.2 | 59 (5.2%) | 41.5 | 80.5 |
| C07 - Beta blocking agents | 36 (1.7%) | 40 (1.9%) | 85 | 35.2 | 31 (1.8%) | 98 | 0 | 35 (3.1%) | 82 | 76 |
| C08 - Calcium channel blockers | 7 (0.3%) | 7 (0.3%) | 85 | 5.5 | 6 (0.3%) | 98 | 0 | 6 (0.5%) | 67 | 73.5 |
| C09 - Agents acting on the renin-angiotensin system | 9 (0.4%) | 1 (0.0%) | 28 | 0 | 0 (0.0%) | NA | NA | 0 (0.0%) | NA | NA |
| C10 - Lipid modifying agents | 6 (0.3%) | 3 (0.1%) | 13.5 | 12 | 0 (0.0%) | NA | NA | 0 (0.0%) | NA | NA |
| D01 - Antifungals for dermatological use | 5 (0.2%) | 16 (0.8%) | 8 | 15.5 | 21 (1.2%) | 9 | 19.5 | 15 (1.3%) | 5 | 8.5 |
| D02 - Emollients and protectives | 3 (0.1%) | 3 (0.1%) | 5 | 18 | 4 (0.2%) | 8.5 | 33.8 | 1 (0.1%) | 103 | 0 |
| D03 - Preparations for treatment of wounds and ulcers | 0 (0.0%) | 0 (0.0%) | NA | NA | 1 (0.1%) | 13.5 | 8.5 | 0 (0.0%) | NA | NA |
| D04 - Antipruritics, including antihistamines, anesthetics, etc. | 0 (0.0%) | 0 (0.0%) | NA | NA | 0 (0.0%) | NA | NA | 1 (0.1%) | 47 | 0 |
| D05 - Antipsoriatics | 2 (0.1%) | 2 (0.1%) | 44 | 41 | 2 (0.1%) | 56.5 | 41.5 | 2 (0.2%) | 14 | 44.5 |
| D06 - Antibiotics and chemotherapeutics for dermatological use | 8 (0.4%) | 11 (0.5%) | 12 | 21.5 | 11 (0.6%) | 4 | 4 | 5 (0.4%) | 4 | 2 |
| D07 - Corticosteroids, dermatological preparations | 30 (1.4%) | 33 (1.6%) | 5.5 | 9.8 | 38 (2.2%) | 7 | 11 | 17 (1.5%) | 8.5 | 39.2 |
| D08 - Antiseptics and disinfectants | 3 (0.1%) | 5 (0.2%) | 9.5 | 11.2 | 4 (0.2%) | 5.5 | 26.2 | 3 (0.3%) | 3 | 45 |
| D10 - Anti-acne preparations | 4 (0.2%) | 3 (0.1%) | 7 | 9.5 | 4 (0.2%) | 10 | 6 | 3 (0.3%) | 10 | 37.5 |
| D11 - Other dermatological preparations | 5 (0.2%) | 5 (0.2%) | 20 | 15 | 4 (0.2%) | 12 | 16 | 1 (0.1%) | 10 | 0 |
| G01 - Gynecological anti-infectives and antiseptics | 10 (0.5%) | 15 (0.7%) | 7 | 8 | 41 (2.3%) | 7 | 3 | 26 (2.3%) | 8 | 2.2 |
| G02 - Other gynecologicals | 95 (4.5%) | 11 (0.5%) | 27.5 | 33.8 | 4 (0.2%) | 74.5 | 59.2 | 12 (1.1%) | 1 | 2 |
| G03 - Sex hormones and modulators of the genital system | 474 (22.6%) | 295 (14.1%) | 38 | 54 | 118 (6.7%) | 13.5 | 47.8 | 53 (4.7%) | 51 | 41 |
| G04 - Urologicals | 1 (0.0%) | 1 (0.0%) | 39 | 0 | 0 (0.0%) | NA | NA | 0 (0.0%) | NA | NA |
| H01 - Pituitary and hypothalamic hormones and analogues | 49 (2.3%) | 28 (1.3%) | 12 | 3 | 2 (0.1%) | 46 | 35.5 | 0 (0.0%) | NA | NA |
| H02 - Corticosteroids for systemic use | 20 (1.0%) | 21 (1.0%) | 14 | 76.5 | 16 (0.9%) | 52 | 96 | 16 (1.4%) | 16.5 | 41 |
| H03 - Thyroid therapy | 157 (7.5%) | 214 (10.2%) | 85 | 38 | 202 (11.4%) | 98 | 7.2 | 145 (12.8%) | 94.5 | 14.8 |
| H04 - Anti-thyroid preparations | 0 (0.0%) | 0 (0.0%) | NA | NA | 0 (0.0%) | NA | NA | 1 (0.1%) | 23 | 0 |
| J01 - Antibacterials for systemic use | 69 (3.3%) | 86 (4.1%) | 7.5 | 5 | 165 (9.3%) | 6.5 | 4 | 90 (7.9%) | 6 | 5 |

| | | | | | | | | | | |
|---|---|---|---|---|---|---|---|---|---|---|
| J02 - Antimycotics for systemic use | 2 (0.1%) | 3 (0.1%) | 1 | 0 | 3 (0.2%) | 1 | 0 | 4 (0.4%) | 1.5 | 1 |
| J05 - Antivirals for systemic use | 9 (0.4%) | 15 (0.7%) | 28.5 | 50.8 | 15 (0.8%) | 43 | 59.2 | 5 (0.4%) | 35 | 33 |
| J06 - Immune sera and immunoglobulins | 3 (0.1%) | 4 (0.2%) | 1 | 0 | 4 (0.2%) | 1 | 0 | 42 (3.7%) | 1 | 0 |
| J07 - Vaccines | 200 (9.5%) | 274 (13.1%) | 1 | 0 | 525 (29.7%) | 1 | 0 | 785 (69.1%) | 1 | 0 |
| L01 - Antineoplastic agents | 4 (0.2%) | 2 (0.1%) | 85 | 0 | 2 (0.1%) | 98 | 0 | 2 (0.2%) | 91 | 4 |
| L02 - Endocrine therapy | 101 (4.8%) | 65 (3.1%) | 8 | 5 | 0 (0.0%) | NA | NA | 0 (0.0%) | NA | NA |
| L04 - Immunosuppressants | 47 (2.2%) | 44 (2.1%) | 6 | 82 | 28 (1.6%) | 6 | 93.5 | 14 (1.2%) | 3 | 10 |
| M01 - Anti-inflammatory and antirheumatic products | 201 (9.6%) | 85 (4.1%) | 9 | 13.8 | 15 (0.8%) | 5 | 10.2 | 1 (0.1%) | 1 | 0 |
| M02 - Topical products for joint and muscular pain | 6 (0.3%) | 6 (0.3%) | 9.5 | 13 | 4 (0.2%) | 1 | 1 | 0 (0.0%) | NA | NA |
| M03 - Muscle relaxants | 2 (0.1%) | 2 (0.1%) | 60.5 | 24.5 | 1 (0.1%) | 98 | 0 | 0 (0.0%) | NA | NA |
| M09 - Other therapeutic products for musculoskeletal system | 0 (0.0%) | 1 (0.0%) | 1 | 0 | 1 (0.1%) | 1 | 0 | 0 (0.0%) | NA | NA |
| N01 - Anesthetics | 7 (0.3%) | 4 (0.2%) | 3 | 18 | 7 (0.4%) | 1 | 0.5 | 6 (0.5%) | 1 | 3 |
| N02 - Analgesics | 808 (38.5%) | 774 (36.9%) | 3 | 4 | 626 (35.4%) | 3 | 3 | 247 (21.7%) | 3 | 5 |
| N03 - Anti-epileptics | 41 (2.0%) | 34 (1.6%) | 85 | 0 | 26 (1.5%) | 98 | 0 | 17 (1.5%) | 98 | 7.5 |
| N04 - Anti-Parkinson drugs | 1 (0.0%) | 0 (0.0%) | NA | NA | 0 (0.0%) | NA | NA | 0 (0.0%) | NA | NA |
| N05 - Psycholeptics | 41 (2.0%) | 34 (1.6%) | 28 | 80 | 22 (1.2%) | 21.5 | 93 | 22 (1.9%) | 28 | 84 |
| N06 - Psychoanaleptics | 141 (6.7%) | 127 (6.1%) | 85 | 19 | 94 (5.3%) | 98 | 0 | 60 (5.3%) | 89 | 15 |
| N07 - Other nervous system drugs | 5 (0.2%) | 6 (0.3%) | 29 | 9.2 | 2 (0.1%) | 66.5 | 31.5 | 1 (0.1%) | 89 | 0 |
| P01 - Antiprotozoals | 14 (0.7%) | 12 (0.6%) | 85 | 63.8 | 8 (0.5%) | 98 | 18 | 2 (0.2%) | 45 | 25 |
| P02 - Anthelmintics | 1 (0.0%) | 4 (0.2%) | 5.5 | 5.8 | 2 (0.1%) | 8 | 7 | 2 (0.2%) | 8.5 | 6.5 |
| P03 - Ectoparasiticides, including scabicides, insecticides | 0 (0.0%) | 0 (0.0%) | NA | NA | 1 (0.1%) | 8 | 0 | 1 (0.1%) | 8 | 0 |
| R01 - Nasal preparations | 177 (8.4%) | 175 (8.3%) | 14 | 80 | 209 (11.8%) | 10 | 40 | 121 (10.7%) | 11 | 59 |
| R02 - Throat preparations | 21 (1.0%) | 41 (2.0%) | 4 | 5 | 43 (2.4%) | 3.5 | 3.8 | 14 (1.2%) | 5.5 | 7.5 |
| R03 - Drugs for obstructive airway diseases | 81 (3.9%) | 83 (4.0%) | 85 | 66 | 80 (4.5%) | 98 | 90 | 53 (4.7%) | 88 | 89.5 |
| R05 - Cough and cold preparations | 16 (0.8%) | 31 (1.5%) | 6 | 9 | 44 (2.5%) | 6 | 3.5 | 17 (1.5%) | 6 | 3 |
| R06 - Antihistamines for systemic use | 267 (12.7%) | 755 (36.0%) | 34 | 27 | 620 (35.1%) | 53 | 78 | 222 (19.5%) | 82 | 76.8 |
| R07 - Other respiratory system products | 1 (0.0%) | 1 (0.0%) | 85 | 0 | 1 (0.1%) | 98 | 0 | 0 (0.0%) | NA | NA |
| S01 - Ophthalmologicals | 11 (0.5%) | 10 (0.5%) | 40 | 81 | 13 (0.7%) | 16 | 37 | 5 (0.4%) | 6 | 91 |
| S02 - Otologicals | 1 (0.0%) | 2 (0.1%) | 6.5 | 1.5 | 4 (0.2%) | 5 | 8 | 1 (0.1%) | 7 | 0 |
| S03 - Other ophthalmologicals and otologicals | 0 (0.0%) | 0 (0.0%) | NA | NA | 5 (0.3%) | 7 | 6 | 2 (0.2%) | 6 | 0 |
| V01 - Allergens | 6 (0.3%) | 5 (0.2%) | 19.5 | 69.8 | 3 (0.2%) | 22.5 | 56.5 | 1 (0.1%) | 92 | 0 |
| V04 - Surgical dressings and sutures | 1 (0.0%) | 0 (0.0%) | NA | NA | 0 (0.0%) | NA | NA | 0 (0.0%) | NA | NA |
| V08 - Contrast media | 1 (0.0%) | 0 (0.0%) | NA | NA | 0 (0.0%) | NA | NA | 0 (0.0%) | NA | NA |

***Supplementary List 1.*** *Medications screened as potentially teratogenic (irrespective of the trimester).*

- Acenocoumarol
- Warfarin
- Fenprocoumon
- Valproate (Valproic acid)
- Topiramate
- Phenobarbital
- Phenytoin
- Isotretinoin
- Acitretin
- Modafinil
- Finasteride
- Methotrexate
- Mycophenolate mofetil
- Tamoxifen
- Thalidomide
- ACE inhibitors
- Angiotensin II receptor blockers (ARBs)
- Carbamazepine
- Non-steroidal anti-inflammatory drugs (NSAIDs)
- Hormonal contraceptives

***Supplementary Table 4.*** *Vaccine exposure by pregnancy trimester and gestational timing.*

| | Preconception (N = 2096) | Trimester 1 (N = 2096) | | Trimester 2 (N = 1767) | | Trimester 3 (N = 1136) | |
|---|---|---|---|---|---|---|---|
| | n (%) | n (%) | Median GA (days) | n (%) | Median GA (days) | n (%) | Median GA (days) |
| Pertussis vaccine | 9 (0.4%) | 2 (0.1%) | 56.5 | 175 (9.9%) | 176 | 650 (57.2%) | 206 |
| Influenza vaccine | 119 (5.7%) | 216 (10.3%) | 54 | 319 (18.1%) | 126 | 149 (13.1%) | 220 |
| COVID-19 vaccine | 102 (4.9%) | 112 (5.3%) | 59 | 182 (10.3%) | 126 | 67 (5.9%) | 216 |
| RSV vaccine (maternal) | 0 (0.0%) | 0 (0.0%) | NA | 2 (0.1%) | 138 | 60 (5.3%) | 237 |
| Hepatitis A vaccine | 8 (0.4%) | 2 (0.1%) | 28.5 | 1 (0.1%) | NA | 0 (0.0%) | NA |
| Hepatitis B vaccine | 0 (0.0%) | 0 (0.0%) | NA | 1 (0.1%) | 123 | 0 (0.0%) | NA |
| Hepatitis A + B combination vaccine | 2 (0.1%) | 0 (0.0%) | NA | 0 (0.0%) | NA | 0 (0.0%) | NA |
| Yellow fever vaccine | 5 (0.2%) | 1 (0%) | 11 | 0 (0.0%) | NA | 0 (0.0%) | NA |
| Rabies vaccine | 2 (0.1%) | 0 (0.0%) | NA | 0 (0.0%) | NA | 0 (0.0%) | NA |
| Tick-borne encephalitis vaccine | 6 (0.3%) | 1 (0%) | 31 | 2 (0.1%) | 155 | 1 (0.1%) | 198 |
| Typhoid vaccine | 1 (0%) | 0 (0.0%) | NA | 0 (0.0%) | NA | 0 (0.0%) | NA |

*All results are shown as absolute numbers (n) and percentages (%); GA: gestational age in days.*

***Supplementary Table 5.*** *Folic acid use among pregnancies following assisted reproductive technology (ART), across pregnancy trimesters and timing of initiation.*

| | **Start ≥28 days before LMP (N=370[1])** | **Start between 28 and 14 days before LMP (N=370[1])** | **Start after 28 days post LMP (N=370[1])** | **Used folic acid concordant to guideline in Belgium (N=370[1])** | **Use in trimester 1 (N = 372)** | | | **Use in trimester 2 (N = 303)** | | | **Use in trimester 3 (N = 193)** | | |
|---|---|---|---|---|---|---|---|---|---|---|---|---|---|
| | n (%) | n (%) | n (%) | n (%) | | Duration (days) | | | Duration (days) | | | Duration (days) | |
| | | | | | n (%) | Median | IQR | n (%) | Median | IQR | n (%) | Median | IQR |
| **Minimum 0.4mg of folic acid** | 291 (78.6%)[2] | 11 (3.0%)[2] | 26 (7.0%)[2] | 274 (74.1%) | 371 (99.7%) | 53 | 48 | 302 (99.7%) | 81 | 57.8 | 193 (100.0%) | 53 | 78 |

*All results are shown as absolute numbers (n) and percentages (%); IQR: interquartile range.[1] Includes all pregnancies that were followed until the end of the first trimester, with exact start and stop dates for folic acid available (N = 12 had missing start/stop data in the T1 cohort).[2] Regarding timing of initiation, the table presents the following clinically relevant categories: 'start ≥28 days before LMP', 'start between 28 and 14 days before LMP', and 'start after 28 days post-LMP'. Categories not shown in the table include: 'start between 13 days before LMP and LMP' (n = 18, 4.9%), 'start after LMP but before 28 days post-LMP' (n = 22, 5.9%), 'pregnancies with a stop date ≥28 days before LMP' (n = 1, 0.3%), and 'pregnancies with no folic acid registrations in T1' (n = 1 , 0.3%)*
